# Unusually high risks of COVID-19 mortality with age-related comorbidities: An adjusted method to improve the risk assessment of mortality using the comorbid mortality data

**DOI:** 10.1101/2021.05.20.21257550

**Authors:** Andrew Antos, Ming Lai Kwong, Timothy Balmorez, Alyssa Villanueva, Shin Murakami

**Affiliations:** Department of Basic Sciences, College of Osteopathic Medicine, Touro University, California, Vallejo, CA

## Abstract

**Background:** The pandemic of Coronavirus Disease 2019 (COVID-19) has been a threat to global health. In the US, the Centers for Disease Control and Prevention (CDC) has listed 12 comorbidities within the first tier that increase with the risk of severe illness from COVID-19, including the comorbidities that are common with increasing age (referred to as age-related comorbidities) and other comorbidities. However, the current method compares a population with and without a particular disease (or disorder), which may result in a bias on the results. Thus, comorbidity risks of COVID-19 mortality may be underestimated.

**Objective:** To re-evaluate the mortality data from US States and estimate the odds ratios of death by major comorbidities with COVID-19, we incorporated the control population with no comorbidity reported and assessed the risk of COVID-19 mortality with comorbidity.

**Methods:** We collected all the comorbidity data from the Public Health websites of fifty US States and Washington DC originally accessed on December 2020. The timing of the data collection should allow minimizing a bias from the COVID-19 vaccines and new COVID-19 variants. The comorbidity demographic data were extracted from the State Public Health data made available online. Using the inverse-variance random-effects model, we performed a comparative analysis and estimated the odds ratio of deaths by COVID-19 with pre-existing comorbidities.

**Results:** A total of 39,451 COVID-19 deaths were identified from four States that had comorbidity data, including Alabama, Louisiana, Mississippi, New York. 92.8% of the COVID-19 deaths were associated with pre-existing comorbidity. The risk of mortality associated with at least one comorbidity combined was 1,113 times higher than that with no comorbidity. The comparative analysis identified nine comorbidities with odds ratios of up to 35 times significantly higher than no comorbidities. Of them, the top four comorbidities were: hypertension (odds ratio 34.73; 95% CI 3.63-331.91; p = 0.002), diabetes (odds ratio 20.16; 95% CI 5.55-73.18; p < 0.00001), cardiovascular disease (odds ratio 18.91; 95% CI 2.88-124.38; p = 0.002); and chronic kidney disease (odds ratio 12.34; 95% CI 9.90-15.39; p < 0.00001). Interestingly, lung disease added only a modest increase in risk (odds ratio 6.69; 95% CI 1.06-42.26; p < 0.00001).

**Conclusion:** The aforementioned comorbidities show surprisingly high risks of COVID-19 mortality when compared to the population with no comorbidity. Major comorbidities were enriched with pre-existing comorbidities that are common with increasing age (cardiovascular disease, diabetes, and hypertension). The COVID-19 deaths were mostly associated with at least one comorbidity, which may be a source of the bias leading to the underestimation of the mortality risks previously reported. We note that the method has limitations stemming primarily from the availability of the data. Taken together, this type of study is useful to approximate the risks, which most likely provide an updated awareness of age-related comorbidities.

## INTRODUCTION

COVID-19 is an infectious disease caused by Severe Acute Respiratory Syndrome Coronavirus-2 (SARS-CoV-2) [1]. It has infected over 82 million people worldwide and over 19 million people in the United States (John Hopkins University Corona Virus Online Map) [2] with cases increasing daily. As of 12/30/2020, over 1.8 million people have died from COVID-19 globally, and 341,313 deaths are currently attributed to COVID-19 in the US [2].

The Centers for Disease Control and Prevention (CDC) has published a list of medical conditions that increase the risk of severe illness from COVID-19 [3]. These comorbid diseases have two tiers; the first tier has 12 conditions that “are (an) increased risk of severe illness from the virus that causes COVID-19”; and the second tier has 11 conditions that “might be at an increased risk for severe illness from the virus that causes COVID-19” [4]. The CDC listed 125 publications for the evidence [4] (updated 11/30/2020; Accessed 3/28/2021). Our rationales are the followings. Firstly, the vast majority of them use data from other countries or mixtures of different countries. The US has a diverse population and COVID-19 has been disproportionately impacted compared to the rest of the world; therefore, the risk of COVID-19 should be adjusted based on the US population. Secondly, the current risk assessment of COVID-19 mortality compares a population with and without a particular disease (or disorder). If the vast majority of the COVID-19 deaths are expected to be with comorbidities, the control population without a particular disease would include the other comorbidities, resulting in a possibility of underestimation of the mortality risks. We reasoned that the control without pre-existing comorbidities should be appropriate as a control population for the mortality risk assessment. To reduce unexpected bias from the incomplete US-State data, we focused on the mortality data as well as incorporated the format of the systematic review. Our objective is to re-evaluate the mortality data from US States and estimate the odds ratios of death by major comorbidities with COVID-19, we incorporated the control population with no comorbidity reported and assessed the risk of COVID-19 mortality with comorbidity. This study provides a new index that provides an overview of the COVID-19 deaths associated with comorbidity.

## METHODS

We followed the Preferred Reporting Items for Systematic Reviews and Meta-Analyses Protocols (PRISMA) statement [5]. Criteria for eligibility is to have comorbidity data made available to the public. The US comorbidity data was searched on PubMed and Scopus for literature search and UpToDate for the clinical overview, using the keywords, “COVID 19 AND Comorbidity”, “COVID 19 AND Comorbidities AND United States”, “COVID 19 AND Comorbidities AND United States AND Mortality”, for example. For US-States and CDC data, fifty State health department, Washington DC websites and the CDC websites were directly accessed; the links of a total of 52 websites searched are shown in Supplementary Material 1. All the data were collected from the websites, links and search functions embedded on the webpages (Originally accessed on June 2020; Updated December 2020). Six states provided comorbidity data: Alabama, Louisiana, Mississippi, New York, Georgia, and Pennsylvania. Pennsylvania was not included in our results because the state department reported incomplete data - noting that 38% of deaths did not include comorbidity data (Pennsylvania Department of Health) [6]. We found data from a study conducted in the State of Georgia, but this study did not list the deaths by comorbidity and thus was not included in this study. Data extraction from reports were made directly from the webpage and duplicated. The webpage links including the States were listed in the Supplementary Material 2. Confirmation of the data was made independently by two investigators.

The Alabama, Louisiana, Mississippi, and New York state department websites update the death tolls for their respective states. We referenced the Alabama [7], Mississippi [8], and New York data [9] which were updated on 12/29/2020. We retrieved Louisiana’s data on 12/17/2020; the Louisiana data were dated on 3/31/2020 [11]. The data surrounding deaths due to COVID-19 in Alabama, Louisiana, Mississippi, and New York were separated and additionally summed for analysis. Deaths that included comorbidity were separated by their respective state. Deaths that did not suffer from comorbidity were also separated by state. To estimate the number of comorbidities in Louisiana, we multiplied the percentages with the total number of deaths since their data only provided a comorbidity percentage. We were unable to input the decimal values from the percentage calculations for Louisiana’s numerical fields. Thus, we rounded up if the tenths decimal place was equal to or greater than five and rounded down if the tenths decimal place was less than five. For New York, we also subtracted the total deaths with comorbidity from the total deaths from COVID-19 to deduce how many COVID-19 deaths did not have comorbidity. The risk of bias was independently tested by two investigators. We used ROBINS-I tool (Risk Of Bias in Non-randomized Studies - of Interventions) [10].

To calculate the odds ratios, we used the software, RevMan5 (Cochrane reviews) [12]. The random-effects method was also used to calculate the odds ratios. The inverse variance method was used since it is a common and simple mode to conduct a meta-analysis. “The inverse variance method is so named because the weight given to each study is chosen to be the inverse of the variance of the effect estimate (i.e. one over the square of its standard error). Thus, larger studies, which have smaller standard errors, are given more weight than smaller studies, which have larger standard errors. The inverse-variance random-effects model has been commonly used in the COVID-19 meta-analysis studies, for example, [13] and [14]. This choice minimizes the imprecision (uncertainty of the pooled effect estimate).” Statistical heterogeneity is measured as I^²^>0. The following interpretations for different heterogeneity percentages were given:0% to 40% “might not be important”, 30% to 60% “may represent moderate heterogeneity”, 50% to 90% “may represent substantial heterogeneity”, and 75% to 100% “is considerable heterogeneity.” The “thresholds for the interpretation of I^²^ can be misleading, since the importance of inconsistency depends on several factors.” These observed factors include the “magnitude and direction of effects” and “the strength of evidence for heterogeneity (e.g., P-value from the Chi-squared test, or a confidence interval for I^²^).”

We organized the ten comorbidities between the states to standardize them for analysis as follows. Hypertension was documented in Mississippi and New York that were included to estimate odds ratios of hypertension. For cardiovascular disease (CVD), New York listed detailed diseases with CVD pathologies (atrial fibrillation, coronary artery disease, and stroke comorbidity) [9] and thus were included under the term CVD. Three states showed the data for the immunocompromised conditions, liver disease, and neurological disease odds ratios; and two states for the chronic kidney disease and obesity odds ratio figures. For chronic kidney diseases (CKD) and renal disease (RD), CKD is a part of Renal Disease (RD), and thus RD data include CKD; nonetheless, we did perform the analysis of CKD. The WHO designates stroke as a neurological disorder [15] and the CDC designates it as cardiovascular or cerebrovascular disease [3]. We followed the CDC designation and thus included stroke data into the cardiovascular disease odds ratios. New York had more detailed designations than other States and we treated the New York data as follows. Dementia data were included into Neurologic Disease, and COPD was included in Lung Disease. In this study, Neurological disease was called Neurologic disease for the consistency of disease designations. Despite the designations by the CDC, the four states also did not distinguish the diabetes types (type 1, type 2, other types of diabetes) and is shown as diabetes.

## RESULTS

We modified the assessment procedure of the COVID-19 mortality data and used the control population with no comorbidity (Methods). We focused on mortality data, since the outcomes from the COVID-19 test results could be influenced by different testing systems and by how they were administered and recorded. For the reason, we excluded the COVID-19 test results. Similarly, we excluded hospitalization that may contribute to a bias of the risk assessment. The US comorbidity data was searched on the PubMed and Scopus websites. The results from queries resulted in a total of 1,484 publications, narrowed down to 169 that included a total of 10 observational studies and case reports. The studies/reports were reviewed and data regarding deaths from patients with and without comorbidity in the US were not found (Accessed June 2020). Thus, the CDC and State websites were directly accessed for provisional comorbidity data; the links were listed in Supplementary Material 1. The PRISMA flow diagram on June 2020 is shown in Figure 1. The websites examined were a total of 52. The CDC had a provisional dataset of “comorbidities and other conditions”; the dataset was dominated by “pneumonia and influenza”, “respiratory failure” and “hypertensive diseases” [16] which were mixed with COVID-19 symptoms, causes of deaths and pre-existing conditions. Thus, we did not use the CDC dataset.

**Figure 1.**
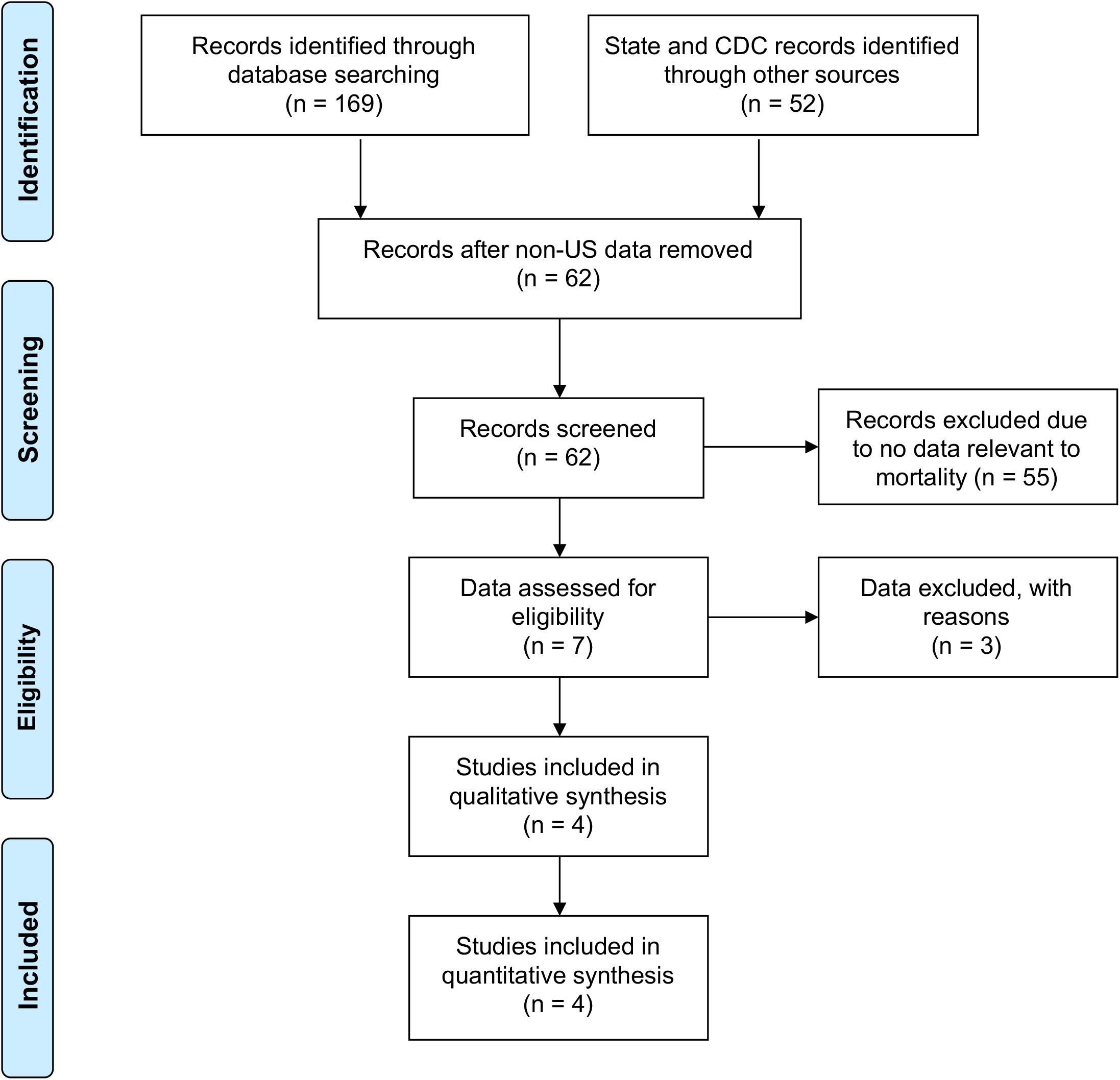
A flow diagram of this study originally accessed on June 2020. The PRISMA template for researchers was used [5].

We collected all the data available which was originally accessed on June 2020 and updated on December 2020 by searching the Public Health websites of fifty States and Washington DC in the US. We have compiled and organized the ten comorbidities between the states to standardize them for analysis (Table 1). The demography is summarized in Supplementary Material 2. The risk of bias within studies was low to modest (Figure 5). More details are described in the Section of Method. A total of 39,451 COVID-19 deaths were found from four states including Alabama, Louisiana, Mississippi, and New York. The results of the total deaths and other separations by the state are summarized in Figure 2. Each of the pre-existing comorbidities associated with COVID-19 is summarized in Figure 3. New York accounts for most of the total COVID-19 deaths in this study, which may create a bias by dominating the result. To minimize the bias, we used the random-effects models to control the results of the odds ratio. As a comparison, New York accounted for 75.4% of the total deaths in the study while Mississippi accounted for 11.9%. Alabama accounted for 12%, while Louisiana accounted for 0.6% of the total deaths in the study. Therefore, the random-effects model helped mitigate some of the bias from New York. 92.8% of the COVID-19 mortality was seen in patients with at least one comorbidity.

**Table 1.** Summary of the odds ratio outcomes.

| Comorbidity Type | OR (95% CI) | P | I<sup>2</sup> | Heterogeneity<br>P |
| --- | --- | --- | --- | --- |
| Cardiovascular Disease | 18.91 (2.88, 124.38) | 0.002 | 100% | < 0.00001 |
| Chronic Kidney Disease | 12.34 (9.90, 15.39) | < 0.00001 | 0% | 0.58 |
| Diabetes | 20.16 (5.55, 73.18) | < 0.00001 | 99% | < 0.00001 |
| Hypertension | 34.73 (3.63, 331.91) | 0.002 | 99% | < 0.00001 |
| Immunocompromised | 6.89 (3.89, 12.20) | < 0.00001 | 81% | 0.006 |
| Liver Disease | 2.04 (3.89, 12.20) | 0.02 | 77% | 0.01 |
| Lung Disease | 6.69 (1.06, 42.26) | 0.04 | 100% | < 0.00001 |
| Neurological Disease | 4.62 (0.55, 38.63) | 0.16 | 99% | < 0.00001 |
| Obesity | 9.00 (1.59, 51.06) | 0.01 | 98% | < 0.00001 |
| Renal Disease | 7.74 (1.33, 45.05) | 0.02 | 100% | < 0.00001 |
| All comorbidities<br>combined | 1113.59 (157.59,<br>7888.28) | < 0.00001 | 100% | < 0.00001 |

Abbreviation: OR (Odds Ratio)

**Figure 2.**
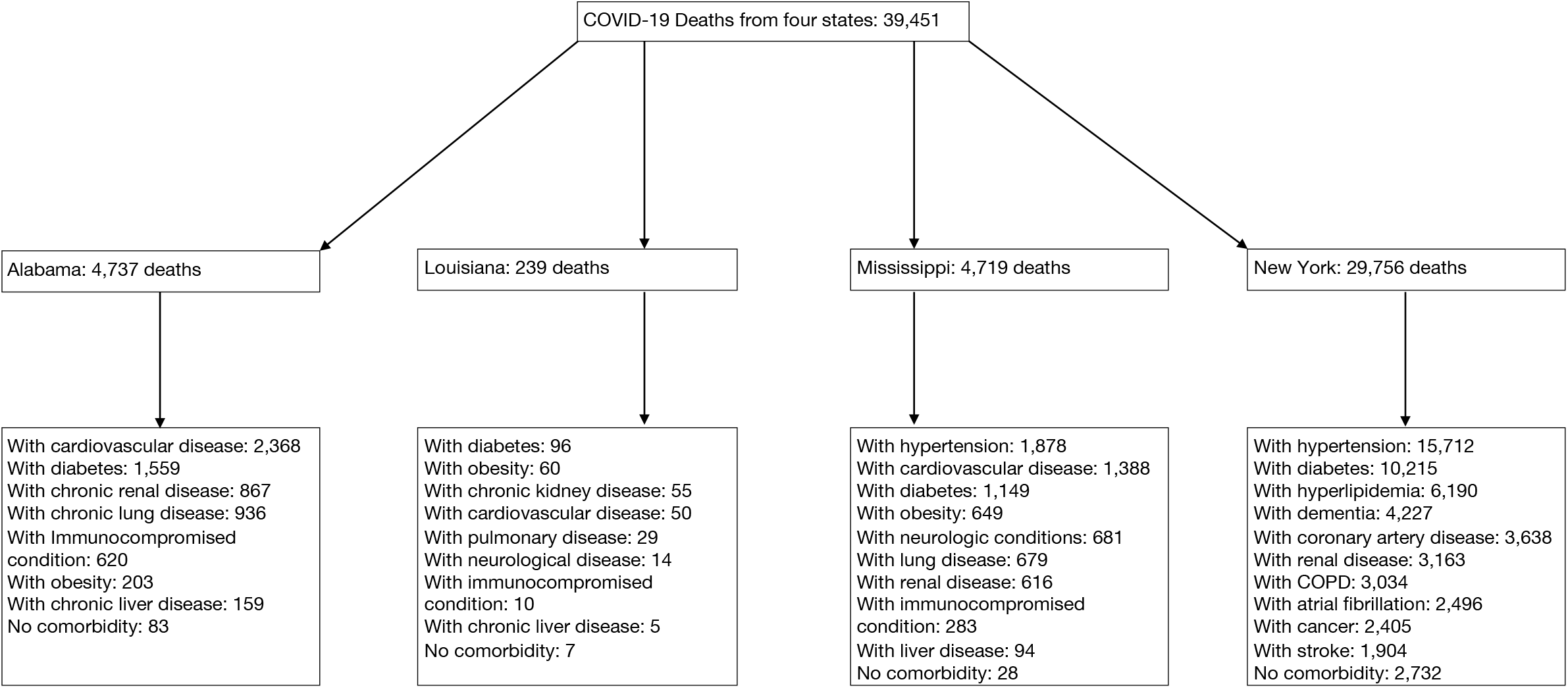
A total number of COVID-19 deaths first separated by state and then by comorbidity. A list of COVID-19 deaths was identified from a total of 4 US states in 2020, including Alabama, Louisiana, Mississippi, and New York.

**Figure 3.**
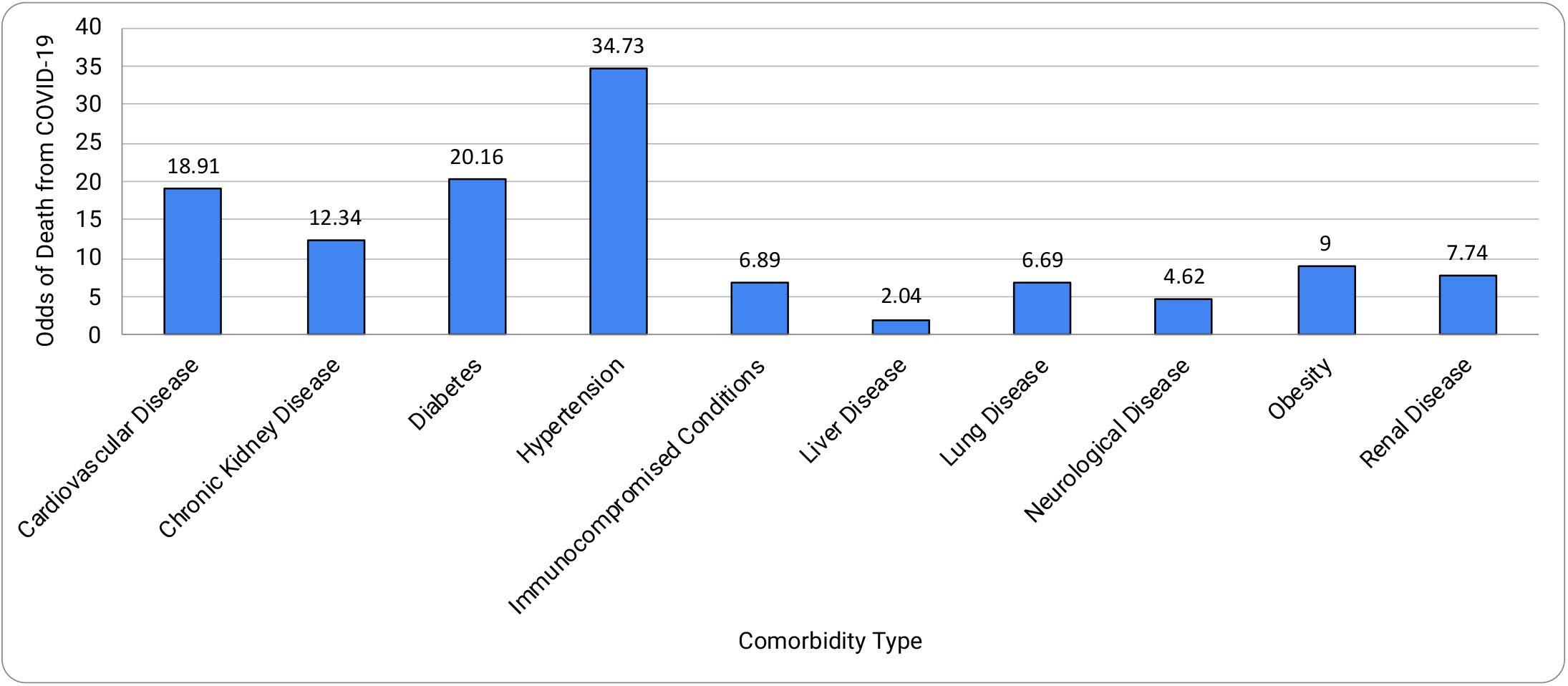
Summary of the mortality risks by COVID-19 comorbidities.

Table 1 and Figure 3 summarize the results. Nine out of ten comorbidities combined to show a surprisingly high risk of death due to covid-19, 1,113 times higher (odds ratio, 1113.59; 95% CI, 157.59, 7888.28; p <.00001). Of all the comorbidities, the top comorbidities were: cardiovascular disease; chronic kidney disease, diabetes; and hypertension. Nine comorbidities, except chronic kidney disease, showed a significant amount of heterogeneity (I^²^ value ranging from 77% to 100%) present which was supported by a strong heterogeneity p-value (ranging from 0.00001 to 0.01) and high variability in the confidence interval (Table 1). The risk of bias across studies were low to modest (Figure 5).

As shown in Figure 4A, cardiovascular disease has a high risk of death due to covid-19 (odds ratio 18.91; 95% CI 2.88-124.38; p = 0.002). Hypertension is often coupled with cardiovascular disease, and when we combined both hypertension and cardiovascular disease, the risk was doubled (odds ratio, 40.70; 95% CI 27.06, 61.21; p < 0.00001). Figure 4B shows the chronic kidney disease (CKD) comorbidity data (odds ratio 12.34; 95% CI 9.90-15.39; p < 0.00001). Importantly, no heterogeneity (I^²^ = 0%) was present in CKD which is supported by the weak P-value (P = 0.58) and significant overlap of confidence intervals between the two studies. Figure 4C shows the diabetes comorbidity data (odds ratio 20.16; 95% CI 5.55-73.18; p < 0.00001). Figure 4D shows the hypertension analysis containing comorbidity data (odds ratio 34.73; 95% CI 3.63-331.91; p = 0.002).

**Figure 4.**
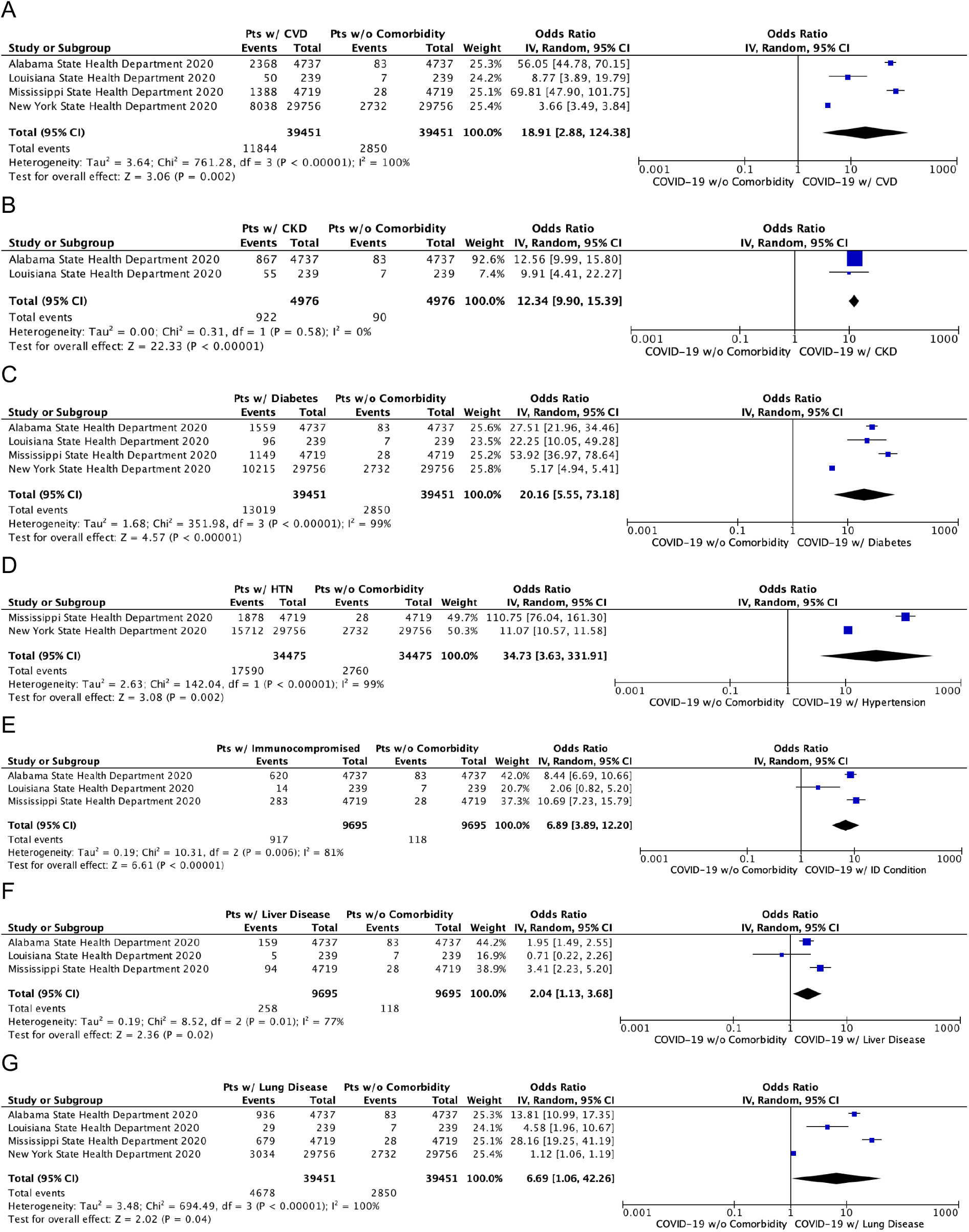
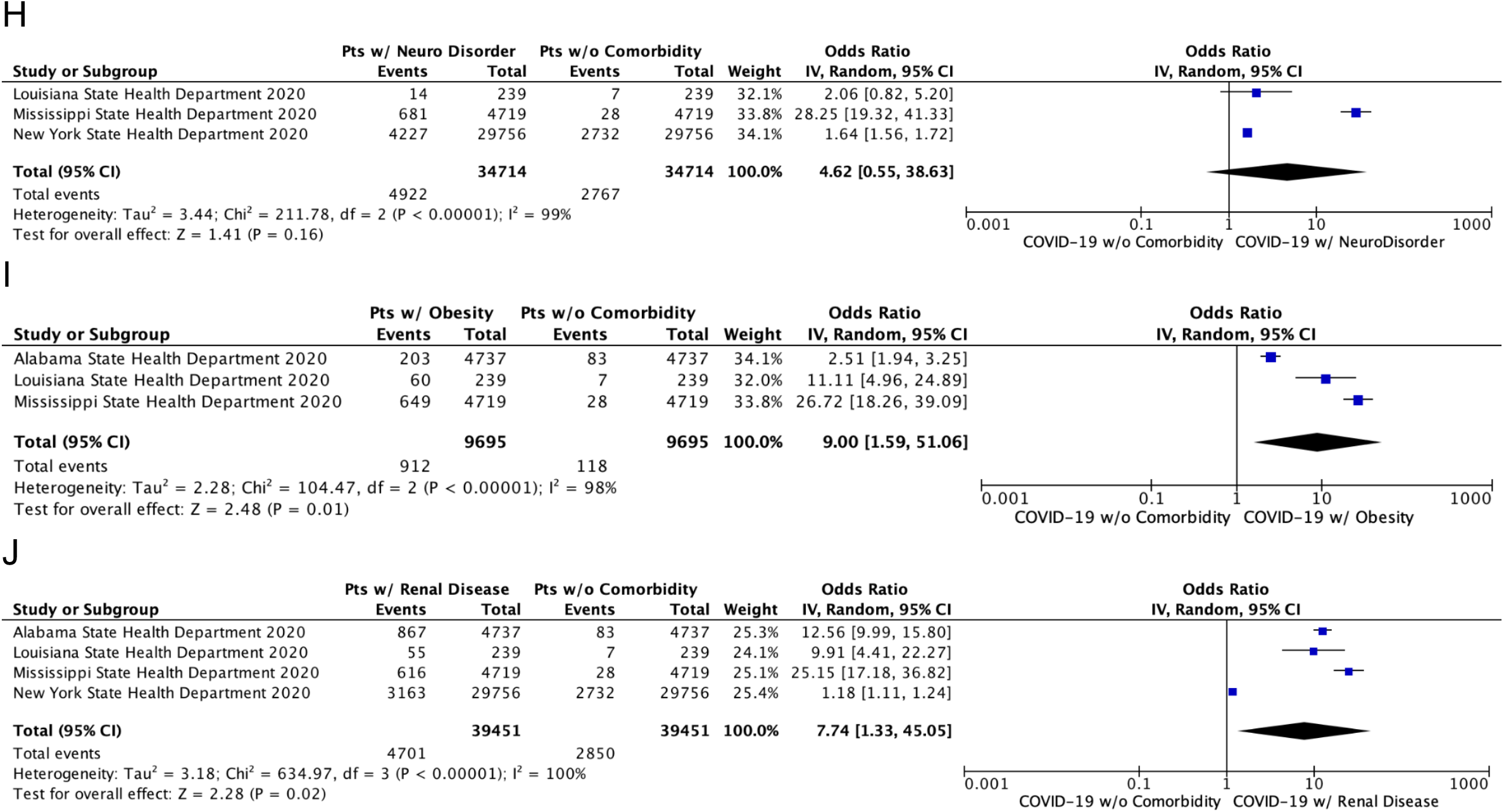
Meta-analysis of the comorbidities with COVID-19. **A**. Cardiovascular disease (CVD). This odds ratio includes atrial fibrillation, coronary artery disease, hypertension, and stroke from New York’s comorbidity data. **B**. Chronic Kidney Disease (CKD). **C**. Diabetes. **D**. Hypertension. **E**. Immunocompromised Condition. **F**. Liver Disease. **G**. Lung Disease. This odds ratio includes Chronic Lung Disease from Alabama’s data and COPD from New York’s data. **H**. Neurological Disease. This odds ratio includes Alzheimer’s disease and Stroke data from New York. **I**. Obesity. **J**. Renal Disease. This odds ratio includes CKD data from Alabama and Louisiana. “Total” columns in the model represent the numerical amount of the total deaths from COVID-The “Events” column on the left represents the number of deaths that included the cardiovascular disease comorbidity in that respective state. The “Events” column on the right represents the number of deaths without comorbidity in that respective state.

**Figure 5.**
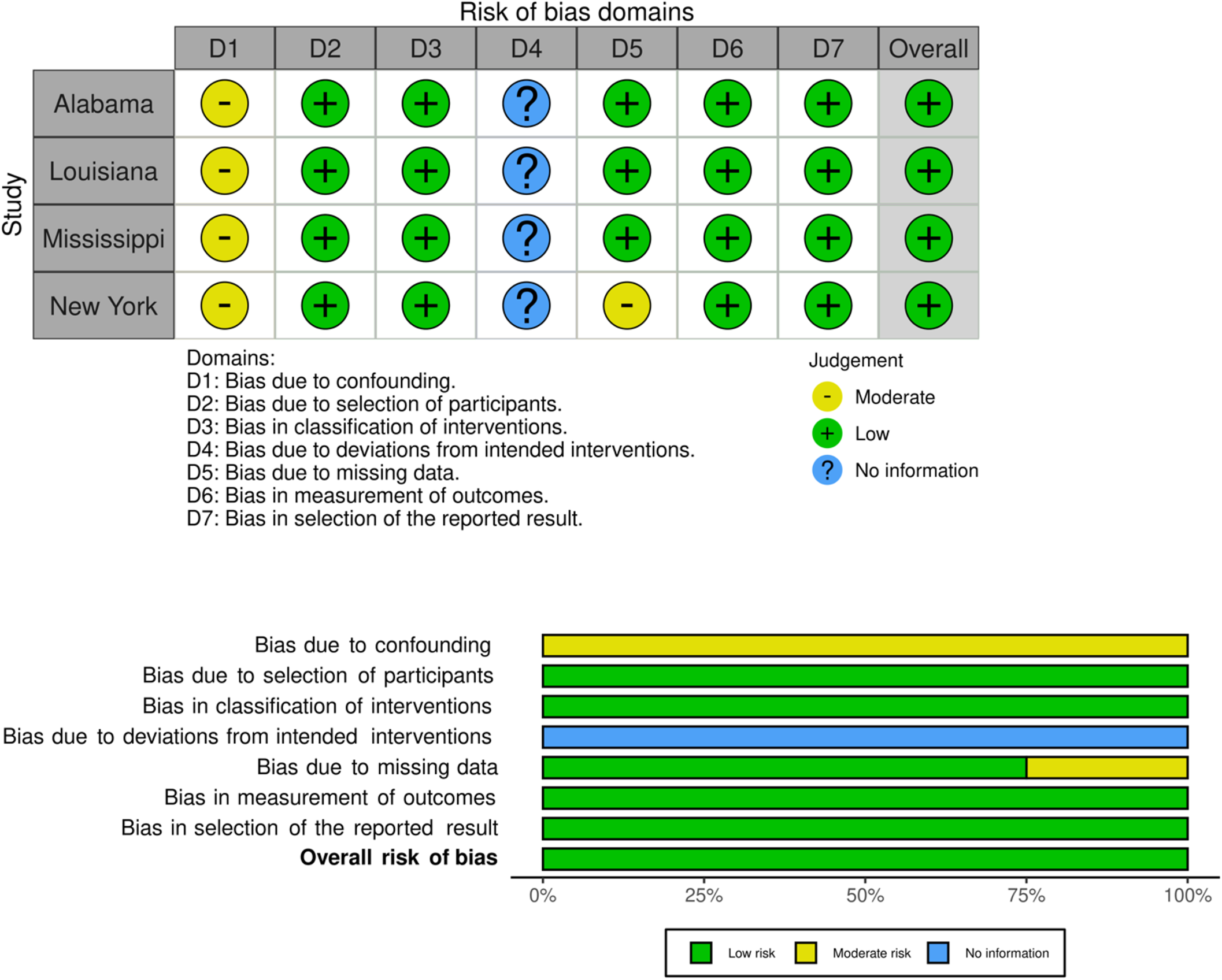
Assessment of the risk of bias using ROBINS-1. Shown are the Traffic light plot (above) and the Summary plot (below). We used comorbidity as an intervention and death as an outcome. No information (Blue) is not applicable to this study. We used a visualization tool as described in [23].

Figure 4E shows the immunocompromised condition comorbidity data (odds ratio, 6.89; 95% CI 3.89, 12.20; p < 0.00001); Figure 4F shows the liver disease comorbidity data (odds ratio, 2.04;95% CI 3.89, 12.20; p = 0.02); and Figure 4G shows the lung disease comorbidity data (odds ratio, 6.69; 95% CI 1.06, 42.26; p = 0.04). The effect of lung disease as a type of comorbidity may have an overlapping, modest effect of death. Neurological condition (Figure 4H) was not significantly different from no comorbidity (odds ratio, 5.41; 95% CI 0.55, 38.63; p = 0.07). It is worth noting that in New York, dementia is one of the leading comorbidities of the COVID-19 deaths, which will require a more specific detailed analysis. Obesity (Figure 4I) had an estimate of 9 (95% CI 1.59, 51.06; p = 0.01). The renal disease (Figure 4J) had an estimate of 7.74 (p = 0.02). A significant amount of heterogeneity (I^²^ > 77%; P< 0.01) was present in this analysis, except for chronic kidney disease (I^²^=0%, heterogeneity P-value = 0.58).

## DISCUSSION

Age-related health conditions including cardiovascular disease, hypertension, and type 2 diabetes are common in the aging population and have been implicated in late-life diseases such as Alzheimer’s disease [17-18]. The centers for disease control and prevention (CDC) have listed comorbidities with the risk of severe illness from COVID-19, including the comorbidities that are common with increasing age (age-related comorbidities) (e.g., heart conditions, hypertension, and type 2 diabetes) and other comorbidities (e.g., smoking, lung disease, and genetic conditions such as down syndrome and sickle cell disease). Most of the previous studies on comorbidities were based on data from other countries. Using the data available from the US, we estimated the odds ratios of comorbidities within the US population to assess their impact on mortality from COVID-19. We found age-related comorbidities to be the major conditions: cardiovascular disease, chronic kidney disease, diabetes, and hypertension. When cardiovascular disease and hypertension were combined, the odds ratio of mortality became 40 times higher than without comorbidity. The risks of mortality from COVID-19 in patients with medical comorbidities appears to be much higher than previous studies that used non-US data. For example, the CDC refers to the studies about hypertension consist most if not all of non-US data (cdc.gov accessed on March 2021). The odds ratios were much higher in the US (odds ratio 34.7; this study) compared to those in studies conducted in non-US-countries (odds ratios 2.3-6.5 in non-US countries) [e.g., 17-20], which implies that there seems to be a variation among countries. Thus, a more comprehensive study remains to be done with data from more US states once they become available.

We organized the comorbidity categories with consideration given to the varied definitions of categories between organizations and state reports. In the study, we followed the CDC categories whenever possible. Adjustments of the categories for the purposes of analysis in this study are described in the Result and Method sections. We also noted that the CDC has a detailed list of diseases contributing to the cause of death, which is dominated by “pneumonia and influenza” and “respiratory failure” in the category [16]. In our analysis, we used pre-existing comorbidities present in the population of the study. We tried to avoid skewed results by using the inverse variance method of data analysis to ensure that the data from New York did not overpower data from the other states. Despite the adjustments, the odds ratios were surprisingly higher than those estimated based on previous research.

Most of the comorbidities showed significantly higher mortality as measured by the odds ratio. Comorbidities that significantly increase the risk of a severe COVID-19 include cardiovascular disease, chronic kidney disease, diabetes, hypertension, immunocompromised condition, liver disease, lung disease, obesity, and renal disease. We could not find a significant increase in risk due to neurological conditions. Heterogeneity was significantly high for nine comorbidities, except for chronic kidney disease. Interestingly, dementia, a part of the neurological conditions, was placed in fourth for comorbidities in New York, while other states did not have this category. Therefore, it remains contested whether or not dementia shows a significant increase in mortality with COVID-19.

We used an adjusted method that has strengths as follows. Firstly, it uses the control population with no comorbidities which avoid bias from the population. The control is particularly important when most of the COVID-19 deaths are associated with one or more comorbidities. Secondly, the mortality data with comorbidities are readily available to the public, which are expected to be less biased. The sample sizes are similar to the previous systematic review and meta-analysis studies [4,19-22]. Thirdly, it is relatively simple to perform the assessment independent of other factors that are expected to have high variability. Finally, we designed the method that avoids the sources of data ambiguity, such as COVID-19 detection systems and variation of the severity of COVID-19. In contrast, the method has limitations stemming primarily from the availability of the data. It is limited to the resources that have comorbidity data made available to the public, though the limitation is similar to that of the meta-analysis. We were also not able to identify the data about more than one comorbidity, which cannot be assessed in the method. Other limitations have been described above (Methods; Results). Since the method focuses on the mortality data, the outcomes from the results gives an overview of the COVID-19 deaths with and without comorbidity. We also note that the comorbidity criteria are inconsistently presented among the States. We propose to have a more unified process of presenting the comorbidities in Public Health data for collaboration purposes.

## CONCLUSIONS

This study is among one of the first US-based studies to show the risk of mortality from COVID-19 in patients who suffer from common comorbidities. We successfully compiled the US COVID-19 mortality data and estimated the risk of death from COVID-19 with and without comorbidity. This type of study is needed to approximate mortality risk in the context of COVID-19 so that we can better stratify how to address and manage patients with these comorbidities from a public health perspective. Nine out of ten comorbidities analyzed were shown to increase the likelihood of death from COVID-19. We found that in the US population, the odds ratio for each of these comorbid conditions was higher than that previously seen in the studies predominantly from outside of the US. We call for a standardized format and distribution of data be provided by each State regarding COVID-19 morbidity and mortality so that a thorough and cohesive analysis of data can occur. We also propose to include a control population with no comorbidity when possible to minimize a bias from other health conditions. Altogether, this study gives an awareness about the comorbidities that are more deleterious than previously recognized and provides a call to action for the public health community.

## Data Availability

All data referred to in the manuscript have been presented in the manuscript.

## Acknowledgment

We thank the members of the Murakami laboratory for useful discussion and technical assistance.

## Supplementary Material 1

The list of the Public Health weblinks of fifty State and the Washington, D.C. websites searched in this study (as of Dec 2020)

1. Alabama: https://www.alabamapublichealth.gov/index.html
2. Alaska: http://hss.state.ak.us/outage.htm
3. Arizona: https://www.azdhs.gov/
4. Arkansas: https://healthy.arkansas.gov/
5. California: https://www.cdph.ca.gov/
6. Colorado: https://cdphe.colorado.gov/
7. Connecticut: https://portal.ct.gov/dph
8. Delaware: https://www.dhss.delaware.gov/dhss/dph/index.html
9. Washington DC: https://dchealth.dc.gov/
10. Florida; http://www.floridahealth.gov/
11. Georgia: https://dph.georgia.gov/
12. Hawaii: https://health.hawaii.gov/
13. Idaho: https://healthandwelfare.idaho.gov/
14. Illinois: https://www.dph.illinois.gov/
15. Indiana: https://www.in.gov/isdh/
16. Iowa: https://idph.iowa.gov/
17. Kansas: https://www.kdheks.gov/
18. Kentucky: https://chfs.ky.gov/agencies/dph/Pages/default.aspx
19. Louisiana: https://ldh.la.gov/
20. Maine: https://www.maine.gov/dhhs/
21. Maryland: https://health.maryland.gov/pages/home.aspx
22. Massachusetts: https://www.mass.gov/orgs/department-of-public-health
23. Michigan: https://www.michigan.gov/mdhhs
24. Minnesota: https://www.health.state.mn.us/
25. Mississippi: https://msdh.ms.gov/
26. Missouri: https://health.mo.gov/index.php
27. Montana: https://dphhs.mt.gov/
28. Nebraska: dhhs.ne.gov/Pages/default.aspx
29. Nevada: http://dpbh.nv.gov/
30. New Hampshire: https://www.dhhs.nh.gov/
31. New Jersey: https://www.nj.gov/health/
32. New Mexico: https://www.nmhealth.org/
33. New York: https://health.ny.gov/
34. North Carolina: https://www.ncdhhs.gov/
35. North Dakota: https://www.health.nd.gov/
36. Ohio: https://odh.ohio.gov/wps/portal/gov/odh/home
37. Oklahoma: https://oklahoma.gov/health.html
38. Oregon: https://www.oregon.gov/oha/ph/pages/index.aspx
39. Pennsylvania: https://www.health.pa.gov/Pages/default.aspx
40. Rhode Island: https://health.ri.gov/
41. South Carolina: https://scdhec.gov/
42. South Dakota: https://doh.sd.gov/
43. Tennessee: https://www.tn.gov/health.html
44. Texas: https://dshs.state.tx.us/
45. Utah: https://health.utah.gov/
46. Vermont: https://www.healthvermont.gov/
47. Virginia: https://www.vdh.virginia.gov/
48. Washington: https://www.doh.wa.gov/
49. West Virginia: https://dhhr.wv.gov/bph/Pages/default.aspx
50. Wisconsin: https://www.dhs.wisconsin.gov/
51. Wyoming: https://health.wyo.gov/

## Supplementary Material 2

### Demography of the population used in this study

| State Health Department | # of Deaths | Gender |  |  | Race/Ethnics |  |  |  |  |  |  | Access Date |
| --- | --- | --- | --- | --- | --- | --- | --- | --- | --- | --- | --- | --- |
|  |  | Male | Female | Unknown | White | Black | Asian | American Indian or Alaska Native | Unknown | Other | Hispanic |  |
| Alabama | 4,737* | 2,487<br>(53%) | 2,240<br>(47%) | 10<br>(0.2%) | 2,723<br>(58%) | 1,544<br>(33%) | 18<br>(0.4%) | NA | 307<br>(7%) | 146<br>(3%) | ** | 12/29/20 |
| Louisiana | 239 | NA | NA | NA | NA | NA | NA | NA | NA | NA | NA | 12/17/20 |
| Mississippi | 4,719 | NA | NA | NA | 2,441 | 1,964 | 15 | 94 | 125 | 80 | ** | 12/29/20 |
| New York | 29,756 | 16,849<br>(57%) | 12,898<br>(43%) | 9<br>(0.0%) | NYC 27%<br>NYS<br>Excluding<br>NYC 66%<br>*** | NYC 28%<br>NYS<br>Excluding<br>NYC 15%<br>*** | NYC 7%<br>NYS<br>Excluding<br>NYC 4%<br>*** | NA | NA | NYC 4%<br>NYS<br>Excluding<br>NYC 3%<br>*** | NYC 34%<br>NYS<br>Excluding<br>NYC 12%<br>*** | 12/29/20 |

Abbreviations: NA (Not available), NYC (New York City), NYS (New York State).
\* The Alabama percentages by race add up more than 100%. It is a possibility of patients in two or more race.
\*\* Alabama and Mississippi label Hispanic as an ethnicity, but not a race. Alabama reports 109 of the total deaths were of Hispanic ethnicity and Mississippi reports 50 deaths were of Hispanic ethnicity.
\*\*\* NYC data and NYS excluding NYC data were available as percentages shown. The numbers were not available.

## REFERENCES

1. Fang, L., Karakiulakis, G., & Roth, M. (2020). Are patients with hypertension and diabetes mellitus at increased risk for COVID-19 infection? The Lancet Respiratory Medicine, 8(4). doi: 10.1016/s2213-2600(20)30116-8 https://www.thelancet.com/journals/lanres/article/PIIS2213-2600(20)30116-8/fulltext

2. Dong E, Du H, Gardner L. (2020) An interactive web-based dashboard to track COVID-19 in real time. Lancet Infect Dis. 20(5):533–534. doi: 10.1016/S1473-3099(20)30120-1. https://www.thelancet.com/journals/laninf/article/PIIS1473-3099(20)30120-1/fulltext

3. The centers for disease control and prevention (CDC). People with Certain Medical Conditions. COVID-19. Retrieved 18 Aug. 2020 from https://www.cdc.gov/coronavirus/2019-ncov/need-extra-precautions/people-with-medical-conditions.html

4. The centers for disease control and prevention (CDC). Underlying Medical Conditions Associated with High Risk for Severe COVID-19: Information for Healthcare Providers Retrieved December 29, 2020 from https://www.cdc.gov/coronavirus/2019-ncov/hcp/clinical-care/underlyingconditions.html

5. Moher D, Liberati A, Tetzlaff J, Altman DG, The PRISMA Group (2009). Preferred Reporting Items for Systematic Reviews and Meta-Analyses: The PRISMA Statement. PLoS Med 6(7): e1000097. doi:10.1371/journal.pmed1000097 https://journals.plos.org/plosmedicine/article?id=10.1371/journal.pmed.1000097

6. Pennsylvania Department of Health. Weekly Report For Deaths Attributed To COVID-19. Retrieved May 17, 2020 from https://www.health.pa.gov/topics/Documents/Diseases%20and%20Conditions/COVID-19%20Death%20Reports/Weekly%20Report%20of%20Deaths%20Attributed%20to%20COVID-19%20--%202020-05-17.pdf?mod=article_inline

7. Alabama Public Health. Data and Surveillance. Retrieved December 29, 2020 rom https://www.alabamapublichealth.gov/covid19/data.html

8. Mississippi State Department of Health. Interactive Charts: COVID-19 Epidemiological Charts and Trends. Retrieved December 29, 2020 from https://msdh.ms.gov/msdhsite/_static/14,21995,420,873.html

9. New York State Department of Health. Workbook: NYS-COVID19-Tracker. (n.d.). Retrieved December 30, 2020 from https://covid19tracker.health.ny.gov/views/NYS-COVID19-Tracker/NYSDOHCOVID-19Tracker-Fatalities?%3Aembed=yes&%3Atoolbar=no&%3Atabs=n

10. Louisiana Department of Health. Updates for 3/31/2020. (n.d.). Retrieved December 17, 2020 from https://ldh.la.gov/index.cfm/newsroom/detail/5522

11. Sterne JAC, Hernán MA, Reeves BC, Savović J, Berkman ND, Viswanathan M, Henry D, Altman DG, Ansari MT, Boutron I, Carpenter JR, Chan AW, Churchill R, Deeks JJ, Hróbjartsson A, Kirkham J, Jüni P, Loke YK, Pigott TD, Ramsay CR, Regidor D, Rothstein HR, Sandhu L, Santaguida PL, Schünemann HJ, Shea B, Shrier I, Tugwell P, Turner L, Valentine JC, Waddington H, Waters E, Wells GA, Whiting PF, Higgins JPT. ROBINS-I: a tool for assessing risk of bias in non-randomized studies of interventions. BMJ 2016; 355; i4919; doi: 10.1136/bmj.i4919. https://www.bmj.com/content/355/bmj.i4919

12. Higgins JPT, Thomas J, Chandler J, Cumpston M, Li T, Page MJ, Welch VA (editors). (2019) Cochrane Handbook for Systematic Reviews of Interventions. 2nd Edition. Chichester (UK): John Wiley & Sons. Cochrane, 2019. www.training.cochrane.org/handbook.

13. Miller LE, Bhattacharyya R, Miller AL. (2020) Diabetes mellitus increases the risk of hospital mortality in patients with Covid-19: Systematic review with meta-analysis. Medicine (Baltimore). 2020 Oct 2;99(40):e22439. doi: 10.1097/MD.0000000000022439. PMID: 33019426 https://www.ncbi.nlm.nih.gov/pmc/articles/PMC7535849/

14. Li J, Yue J, Zhang S, Wu J, Lian R, Zhang R, Cheng P. Medicine (Baltimore). (2020) Relationship between digestive diseases and COVID-19 severity and mortality: A protocol for systematic review and meta-analysis. 99(48):e23353. doi: 10.1097/MD.0000000000023353. PMID: 33235103 https://www.ncbi.nlm.nih.gov/pmc/articles/PMC7710250/

15. World Health Organization (WHO). (2016) What are neurological disorders and how many people are affected by them? (n.d.). Retrieved from https://www.who.int/news-room/q-a-detail/what-are-neurological-disorders

16. The centers for disease control and prevention (CDC). Weekly Updates by Select Demographic and Geographic Characteristics. Retrieved March 31, 2021 from https://www.cdc.gov/nchs/nvss/vsrr/covid_weekly/index.htm

17. Vahdati Nia B, Kang C, Tran MG, Lee D, Murakami S. (2017) Meta Analysis of Human AlzGene Database: Benefits and Limitations of Using C. elegans for the Study of Alzheimer’s Disease and Co-morbid Conditions. Front Genet. 12;8:55. doi: 10.3389/fgene.2017.00055. https://doi.org/10.3389/fgene.2017.00055

18. Le D, Crouch N, Villanueva A, Ta P, Dmitriyev R, et al. 2020. Evidence-Based Genetics and Identification of Key Human Alzheimer’s Disease Alleles with Co-morbidities. J Neurol Exp Neurosci 6(1): 20–24. https://touroscholar.touro.edu/tuccom_pubs/139/

19. Matsushita K, Ding N, Kou M, Hu X, Chen M, Gao Y, Honda Y, Zhao D, Dowdy D, Mok Y, Ishigami J, Appel LJ. 2020 The Relationship of COVID-19 Severity with Cardiovascular Disease and Its Traditional Risk Factors: A Systematic Review and Meta-Analysis. Glob Heart, 15(1):64. doi: 10.5334/gh.814 https://globalheartjournal.com/article/10.5334/gh.814/

20. Guo, X., Y. Zhu, and Y. Hong. 2020 Decreased Mortality of COVID-19 With Renin-Angiotensin-Aldosterone System Inhibitors Therapy in Patients with Hypertension: A Meta-Analysis. Hypertension, 76(2):e13–e14. doi: 10.1161/HYPERTENSIONAHA.120.15572 https://www.ahajournals.org/doi/10.1161/HYPERTENSIONAHA.120.15572

21. Wu T, Zuo Z, Kang S, Jiang L, Luo X, Xia Z, Liu J, Xiao X, Ye M, Deng M. 2020 Multi-organ Dysfunction in Patients with COVID-19: A Systematic Review and Meta-analysis. Aging Dis, 11(4): 874–894. doi: 10.14336/AD.2020.0520 http://www.aginganddisease.org/EN/10.14336/AD.2020.0520

22. Zhang J, Wu J, Sun X, Xue H, Shao J, Cai W, Jing Y, Yue M, Dong C. 2020 Association of hypertension with the severity and fatality of SARS-CoV-2 infection: A meta-analysis. Epidemiol Infect 148:e106. doi: 10.1017/S095026882000117X https://www.cambridge.org/core/journals/epidemiology-and-infection/article/association-of-hypertension-with-the-severity-and-fatality-of-sarscov2-infection-a-metaanalysis/4116FAD7D866737099F976E7E7FAEB15

23. McGuinness, LA, Higgins, JPT. 2020 Risk-of-bias VISualization (robvis): An R package and Shiny web app for visualizing risk-of-bias assessments. Res Syn Meth. 1–7. https://doi.org/10.1002/jrsm.1411

